# A Non-Invasive Ultrasound-Based Model for Assessing Endometrial Cancer Risk in Postmenopausal Bleeding

**DOI:** 10.1101/2025.02.17.25322434

**Authors:** Nagma Khatoon, Zehra Mohsin, Shagufta Wahab, Kafil Akhtar

## Abstract

**Aims:** This study evaluates the risk of endometrial carcinoma in postmenopausal women presenting with bleeding using an ultrasound-based risk scoring system. It aims to integrate imaging parameters and clinical variables for improved differentiation between benign and malignant endometrial conditions.

**Methods:** A prospective analysis was conducted on postmenopausal women presenting with bleeding per vagina. Clinical variables including age, parity, BMI, hypertension, and diabetes were combined with ultrasound findings such as endometrial thickness, echogenicity, endo-myometrial interface, vascular patterns and Doppler scores. A comprehensive scoring system, based on International Endometrial Tumor Analysis (IETA) terminology, was employed to stratify patients’ risk levels. Diagnostic accuracy was determined by comparing the scoring system outcomes with histopathological findings.

**Results:** The ultrasound-based scoring system demonstrated significant diagnostic accuracy in differentiating between benign and malignant endometrial pathologies. Endometrial thickness greater than 4 mm, heterogeneous echogenicity, irregular endo-myometrial interface and increase vascular flow were identified as strong predictors of malignancy. The model achieved a high sensitivity and specificity, providing a non-invasive yet reliable tool for risk stratification in clinical settings. The findings emphasize the potential of an integrated ultrasound-based scoring system in guiding clinical decisions for postmenopausal women with bleeding.

**Conclusion:** We concluded that by incorporating both clinical and imaging parameters improves diagnostic accuracy, minimizes unnecessary invasive procedures, and aids in timely intervention.

## 1. Introduction

Endometrial carcinoma is the most prevalent gynecological malignancy in developed nations and ranks as the sixth most common cancer affecting the female reproductive system globally[1]. Endometrial carcinoma primarily affects postmenopausal women, typically occurring in their sixth and seventh decades of life, with an age incidence ranging from 55 to 70 years [2]. The mean age of patients at the diagnosis of endometrial cancer in developing countries like India is 54 years. The majority of these patients fall within the age group of 40–70 years [3].

Postmenopausal bleeding (PMB) is a key clinical symptom of endometrial carcinoma, with approximately 90% of cases presenting with abnormal uterine bleeding [4]. However, PMB also arises from a spectrum of benign conditions, including endometrial atrophy and polyps, making accurate and timely diagnosis critical [5]. Transvaginal ultrasound (TVUS) is widely employed as the initial diagnostic modality due to its accessibility, cost-effectiveness, and high sensitivity. It allows for the measurement of endometrial thickness, although the optimal cutoff value for determining further management remains uncertain [6]. Measurement of endometrial thickness along with other sonographic features such as echogenicity, vascular patterns, the endo-myometrial interface and Doppler score, offers valuable insights into the likelihood of malignancy[7].

Despite the utility of TVUS, the lack of a definitive endometrial thickness threshold for excluding malignancy underscores the need for an integrated diagnostic approach [8]. Risk-scoring models incorporating both clinical and imaging parameters have demonstrated promise in enhancing diagnostic accuracy and improving risk stratification in women with PMB [9,10].

This study aims to evaluate an ultrasound-based risk-scoring model for symptomatic postmenopausal bleeding women using International Endometrial Tumor Analysis (IETA) terminology including endometrial thickness > 4mm, endometrial echogenicity, Endo-myometrial interface, Doppler Score, vascular pattern and patient characteristics including age, parity, BMI, hypertension, diabetes, hepatic disease to differentiate between benign and malignant endometrium. This approach suggests a comprehensive evaluation method that incorporates different imaging techniques to enhance diagnostic accuracy and guiding subsequent management decision.

## 2. Materials and Methods

This prospective, cross-sectional, observational study included 50 postmenopausal women presenting with vaginal bleeding, evaluated at the Department of Obstetrics and Gynecology in collaboration with Department of Radiology and Department of Pathology, Jawaharlal Nehru Medical College, Aligarh Muslim University from August 2022 to August 2024. Eligibility criteria included postmenopausal women presenting with vaginal bleeding and an Endometrial thickness (ET) of ≥4 mm on Transvaginal sonography (TVS). Exclusion criteria included women with known endometrial malignancy, active pelvic infections, Premenopausal or perimenopausal females, those taking medications which alter the endometrium (HRT, OCP, Tamoxifen, SERM’s), individuals with coagulation and bleeding disorders, those on anticoagulant therapy, patients with cervical abnormalities and those with thyroid disorders. The study was approved by Institutional Ethics Committee. Participants were recruited following informed written consent, ensuring voluntary participation and adherence to ethical guidelines. A comprehensive clinical evaluation was conducted, including a detailed medical history and physical examination (general, Abdominal and pelvic). Clinical data such as age, BMI, parity, and comorbidities (e.g., hypertension, diabetes) were recorded. Lab investigations (CBC, RFT, LFT, PT-INR, Thyroid profile) were done.

High-resolution standard B-mode transvaginal ultrasound (TVUS) with color Doppler was conducted using a Samsung V8 ultrasound machine equipped with a 2-10 MHz transducer, with all scans performed by the same operator. The assessed parameters included endometrial thickness (cutoff value ≥4 mm), endometrial vascularity evaluated using the International Endometrial Tumor Analysis (IETA) scoring system (score 1: no color flow signals, score 2: minimal color flow, score 3: moderate color flow, and score 4: abundant color flow), and vascular pattern classified according to IETA terminology as no vessel, single vessel, multiple dominant vessels, scattered vessels, or circular flow. Additionally, endometrial echogenicity (homogeneous or heterogeneous) and the integrity of the endometrial-myometrial interface (well-defined or ill-defined) were assessed following IETA guidelines.

Following ultrasound evaluation, all participants underwent an endometrial biopsy via dilation and curettage (D&C), Pipelle biopsy, or hysteroscopic-guided biopsy. Histopathological examination was performed to confirm the final diagnosis, and Ultrasound findings were compared with the final histopathology as the gold standard diagnostic test then submitted to Statistical analysis.

## 3. Statistical Analysis

The diagnostic accuracy of the ultrasound-based risk scoring system was evaluated by comparing its results with histopathological findings. Statistical analyses were performed using GraphPad Prism version 9 and IBM SPSS Statistics version 27. Continuous variables were expressed as means ± standard deviations, while categorical variables were presented as frequencies and percentages. Data analysis included a univariate study, with Chi-square tests used for categorical comparisons and t-tests or Mann-Whitney U tests applied for continuous variables. To assess the validity of different ultrasound variables, sensitivity, specificity, positive predictive value (PPV), and negative predictive value (NPV) were calculated. The odds ratio (OR) was used to estimate the risk of malignancy associated with significant variables, then we calculate the risk of malignancy associated with ultrasound parameters, followed by risk scoring for each case. Receiver Operating Characteristic (ROC) curves were generated to determine the optimal cutoff scores for predicting malignancy. A p-value of less than 0.05 was considered statistically significant.

## 4. Results

The study involved 50 postmenopausal women experiencing vaginal bleeding with endometrial thickness equal to or more than 4mm. Patient clinical variables, ultrasound findings and color Doppler assessment of the endometrium were compared to the histopathological results to identify the best predictors of endometrial cancer. Histopathology confirmed endometrial cancer in 16% (n=8) of participants, while 84% (n=42) had benign lesions. Among the benign cases, the most common findings were atrophic endometritis (28%, n=14), endometrial hyperplasia without atypia (24%, n=12), endometrial polyps (10%, n=5), and disordered proliferative endometrium (8%, n=4). Less frequent findings included endometrial hyperplasia with atypia (6%, n=3), chronic tubercular granulomatous endometritis (4%, n=2), luteal phase defect (2%, n=1), and insufficient samples (2%, n=1). (Table 1)

**Table No.1.** Distribution of patients with PMB according to histopathology finding.

| **Histopathology** | **Number of Patients (n)** | **Percentage (%)** |
| --- | --- | --- |
| Atrophic Endometritis | 14 | 28 |
| Endometrial Hyperplasia without atypia | 12 | 24 |
| Endometrial Cancer | 8 | 16 |
| Endometrial Polyp | 5 | 10 |
| Disordered Proliferative Endometrium | 4 | 8 |
| Endometrial Hyperplasia with atypia | 3 | 6 |
| Chronic Tubercular Granulomatous Endometritis | 2 | 4 |
| Luteal Phase Defect | 1 | 2 |
| Insufficient Sample | 1 | 2 |
| Total (n) | 50 | 100% |

Analysis of clinical variables such as age, parity, BMI, hypertension, and liver disease showed no statistically significant differences between benign and malignant cases.(Table 2) However, hypertension, diabetes, and liver disease were associated with slightly higher odds of endometrial cancer, though these findings did not reach statistical significance. (Table 3)(figure 1)

**Table No.2.** Comparison of Clinical Parameters Between Endometrial Cancer and Benign Lesion Groups.

| Parameter | Endometrial Cancer (n=8) | Benign Lesion (n=42) | P Value | 95% confidence interval |
| --- | --- | --- | --- | --- |
| Age (yrs) (Mean) | 57.25 | 55.26 | 0.5676 | -9.487 to 5.510 |
| Parity (Mean) | 4.0 | 3.381 | 0.3612 | -2.068 to 0.8296 |
| BMI (kg/m<sup>2</sup>)(Mean) | 31.04 | 30.46 | 0.6978 | -3.807 to 2.655 |

**Table No.3.** Association of Comorbidities with Endometrial Cancer and Benign Lesions.

| Parameter | Endometrial Cancer (n) | Benign Lesion (n) | P Value | Odds Ratio |
| --- | --- | --- | --- | --- |
| HTN | 5 | 18 | 0.3069 | 2.222 |
| Diabetes | 6 | 16 | 0.0539 | 4.875 |
| Liver Disease | 2 | 6 | 0.4487 | 2.000 |

**Figure 1:**
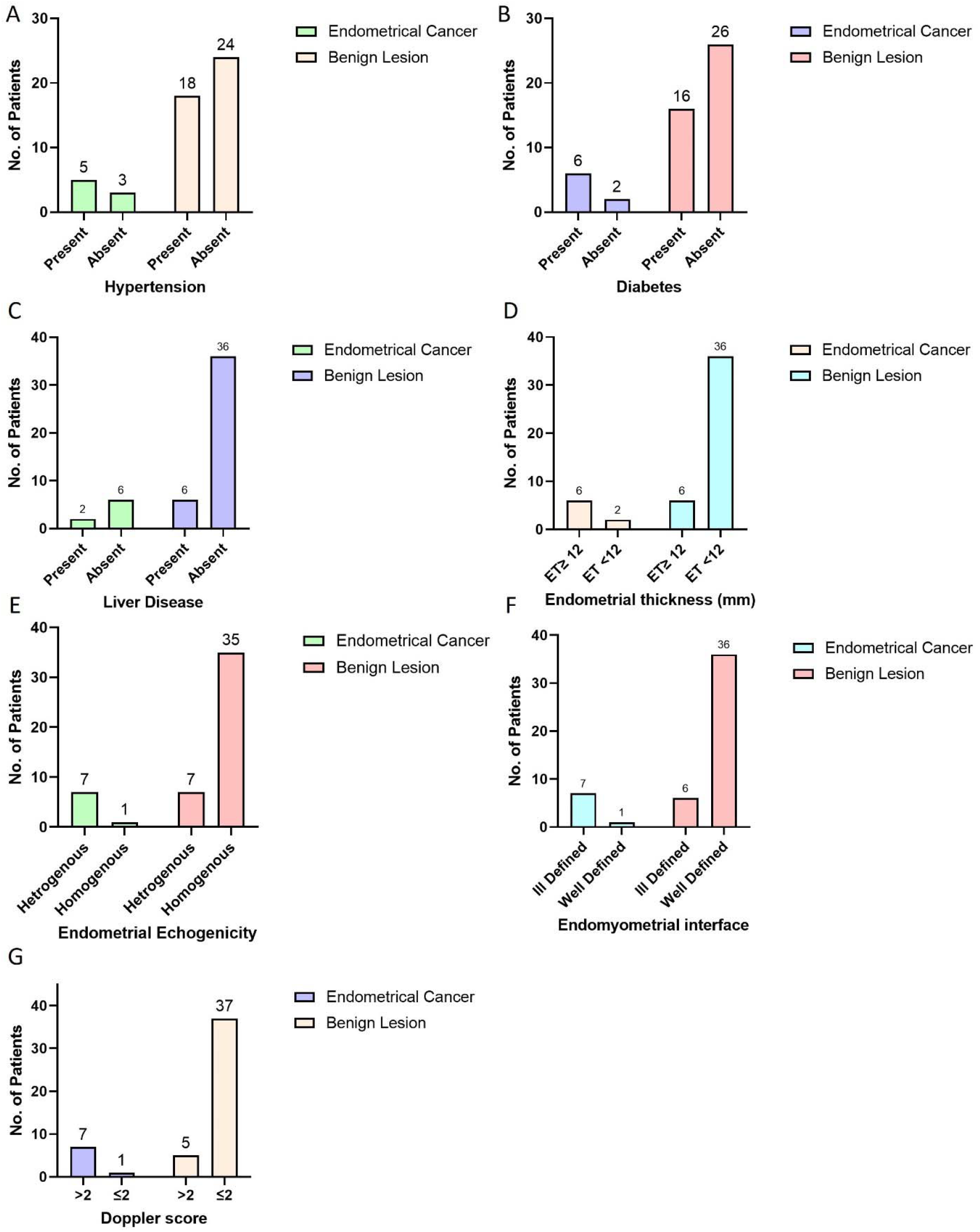
Illustrates the distribution of Clinical and Ultrasound Parameters in Endometrial Cancer and Benign Lesion Groups in the study population.

**Figure 2:**
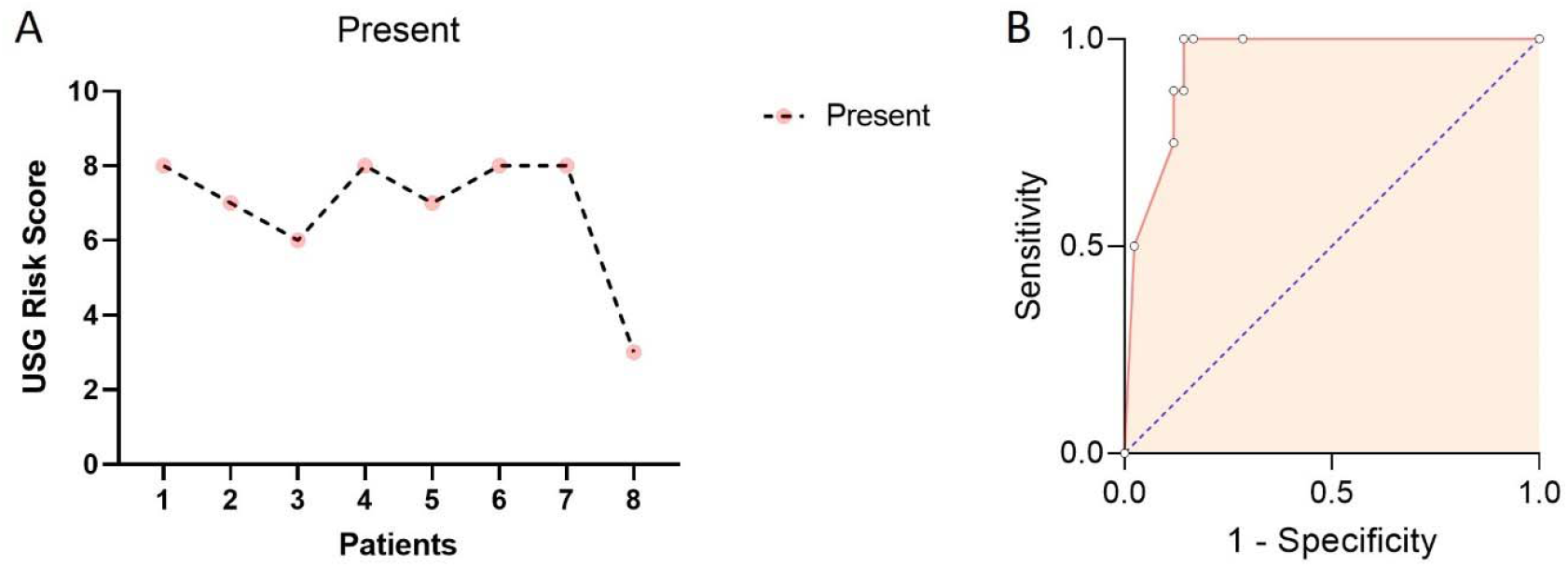
Illustrates the diagnostic Performance of Ultrasound Risk Score in Predicting Endometrial Cancer. Panel A: Distribution of Ultrasound Risk Scores in Patients with Endometrial Cancer. Panel B: Receiver operator curve for estimation of the best cutoff score predicting endometrial cancer (ROC curve).

The ultrasound variables commonly linked to endometrial cancer included an increased endometrial thickness (mean, 14.30 ± 5.14 mm), heterogeneous endometrial echogenicity (87.5%), an ill-defined endo-myometrial interface (87.5%), and a Doppler score > 2 (87.5%). These parameters showed significant differences between benign and malignant cases (p < 0.0001).(table 4) Based on the analysis of significant clinical and ultrasound variables, as determined by a specific ROC curve, the following were identified as strong predictors of malignancy: endometrial thickness ≥12 mm (OR = 35.00), heterogeneous endometrial echogenicity (OR = 35.00), ill-defined endo-myometrial interface (OR = 42.00), and Doppler score > 2 (OR = 51.80).(table 5) These variables exhibited a powerful and significant difference in likelihood ratio results and were selected for inclusion in the designed model. The odds ratio for each ultrasound variable was calculated to estimate the risk of malignancy based on its individual score.(table 6) These predictors were integrated into a composite risk-scoring model, which demonstrated excellent discriminatory performance, with an AUC of 0.943 (95% CI: 0.8813–1.000). The optimal cutoff score of 5.0 yielded a sensitivity of 87.50%, specificity of 88.10%, and balanced diagnostic accuracy, along with an improved likelihood ratio.(table 7)

**Table No.4.** Comparison of Endometrial Ultrasound Characteristics Between Endometrial Cancer and Benign Lesion Groups.

| Parameter | Endometrial Cancer (n) | Benign Lesion (n) | P Value |
| --- | --- | --- | --- |
| Endometrial Thickness (mm) |  |  |  |
| ET>12 | 6 | 6 | 0.0002 |
| ET<12 | 2 | 36 |  |
| Endometrial Echogenicity |  |  |  |
| Heterogenous | 7 | 7 | < 0.0001 |
| Homogenous | 1 | 35 |  |
| Endo myometrial Interface |  |  |  |
| Ill defined | 7 | 6 | < 0.0001 |
| Well Defined | 1 | 36 |  |
| Doppler Score |  |  |  |
| >2 | 7 | 6 | < 0.0001 |
| <2 | 1 | 36 |  |

**Table No.5.** Diagnostic Performance of Ultrasound Parameters in Differentiating Endometrial Cancer from Benign Lesions.

| Parameter | Sensitivity | Specificity | PPV | NPV | OR | P-value |
| --- | --- | --- | --- | --- | --- | --- |
| Endometrial Thickness | 75.00% | 85.71% | 50.00 | 94.74% | 18.00 | 0.0002 |
| Endometrial Echogenicity | 87.50% | 83.33% | 50.00 | 97.22 | 35.00 | <0.0001 |
| Endo myometrial Interface | 87.50% | 85.71% | 53.85 | 97.30 | 42.00 | <0.0001 |
| Doppler Score ($>2$) | 87.50% | 88.10% | 58.33 | 97.37 | 51.80 | <0.0001 |

**Table 6.** Analysis estimating; odds ratio, and the final score for the significant variables.

| Variable | Cut off | Odds | Score | CI | P value |
| --- | --- | --- | --- | --- | --- |
| Endomyometrial Interface | - | 42 | 2 | 4.911 to 479.8 | <0.0001 |
| Endometrial Echogenicity | - | 35 | 2 | 4.279 to 400.7 | <0.0001 |
| Doppler Score | $>2$ | 51.8 | 3 | 5.707 to 590.2 | <0.0001 |
| Endometrial Thickness | $\geq 12$ mm | 18 | 1 | 2.975 to 94.54 | 0.0002 |

**Table 7.** Validity of risk scoring model for the studied population (cutoff score thresholds).

| Score | Sensitivity | 95% CI | Specificity | 95% CI | Likelihood ratio |
| --- | --- | --- | --- | --- | --- |
| > 0.5000 | 1.000 | 0.6756 to 1.000 | 0.7143 | 0.5643 to 0.8283 | 3.500 |
| > 1.500 | 1.000 | 0.6756 to 1.000 | 0.8333 | 0.6940 to 0.9168 | 6.000 |
| > 2.500 | 1.000 | 0.6756 to 1.000 | 0.8571 | 0.7216 to 0.9328 | 7.000 |
| > 3.500 | 0.8750 | 0.5291 to 0.9936 | 0.8571 | 0.7216 to 0.9328 | 6.125 |
| > 5.000 | 0.8750 | 0.5291 to 0.9936 | 0.8810 | 0.7500 to 0.9481 | 7.350 |
| > 6.500 | 0.7500 | 0.4093 to 0.9556 | 0.8810 | 0.7500 to 0.9481 | 6.300 |
| > 7.500 | 0.5000 | 0.2152 to 0.7848 | 0.9762 | 0.8768 to 0.9988 | 21.00 |

## 5. Discussion

Postmenopausal bleeding (PMB) is a significant clinical concern, particularly due to its strong association with endometrial cancer. While the majority of PMB cases stem from benign conditions, approximately 10% of cases are attributed to malignancy, necessitating accurate and early diagnostic methods. This study underscores the critical role of an integrated ultrasound-based risk-scoring system for improving diagnostic precision in differentiating between benign and malignant endometrial conditions postmenopausal women.

A prospective, cross-sectional study was conducted on 50 postmenopausal women presenting with vaginal bleeding and endometrial thickness (ET) of ≥4 mm on Transvaginal sonography (TVS) to test sensitivity of certain clinical and ultrasound variables for prediction of endometrial cancer.

In the initial phase of the analysis, the significance of both clinical and ultrasound variables was assessed individually through univariate analysis. Clinical variables—age, parity, BMI, HPN, and diabetes mellitus—showed no significant difference between benign and malignant cases. In contrast, ultrasound variables, including thickened endometrium (mean 14.30 ± 5.14 mm), heterogeneous echogenicity, ill-defined endometrium-myometrium interface, and Doppler score > 2, demonstrated statistically significant differences between benign and malignant cases. Based on this analysis ultrasound variables were were selected for inclusion in the designed model.

N.M. Madkour [11] conducted a study, developed an ultrasound risk-scoring model for endometrial malignancy, incorporating five key variables: thick endometrium, heterogeneous echogenicity, irregular endometrial midline, ill-defined endo-myometrial interface and Doppler score > 2. ROC curve analysis identified endometrial thickness ≥12 mm (OR = 1.9), heterogeneous endometrium (OR = 2.7), irregular midline (OR = 1.9), ill-defined interface (OR = 3.3), and Doppler score > 2 (OR = 2.5) as significant predictors, with an optimal cutoff score of ≥10 demonstrating 95% accuracy, 80% sensitivity, 97.8% specificity, and an AUC of 0.95. In the present study, four ultrasound variables—endometrial thickness ≥12 mm, heterogeneous endometrial echogenicity, ill-defined endo-myometrial interface, and Doppler score > 2—were identified as the most powerful predictors in the risk-scoring model. These variables exhibited significantly higher odds ratios (endometrial thickness ≥12 mm: OR = 35.00; heterogeneous echogenicity: OR = 35.00; ill-defined interface: OR = 42.00; Doppler score > 2: OR = 51.80) compared to Madkour’s study. The optimal cutoff score of 5.0 yielded a sensitivity of 87.50%, specificity of 88.10%, and an AUC of 0.943 (95% CI: 0.8813–1.000), indicating a more robust predictive model with enhanced diagnostic accuracy. Both studies underscore the critical role of ultrasound variables in predicting endometrial cancer, but the current study demonstrates improved predictive performance, suggesting advancements in diagnostic precision.

Although the odds ratio for endometrial thickness (OR = 18.00, 95% CI: 2.975 to 94.54) with sensitivity of 75% and specificity of 85.71% in the current study (using ET≥ 12 as threshold) was lower than for other variables, it remains a significant predictor of malignancy (p=0.0002), consistent with the established literature. While Seckin et al. (2016)[12] suggested an optimal threshold of 8.2 mm, Long et al. (2020) [13] proposed a lower 5 mm cutoff, and Wong et al. (2016) [14] recommended 3 mm, each reflecting different balances of sensitivity and specificity. Madkour (2017) [11] identified a 12 mm threshold with high sensitivity (90%) and specificity (78.3%) and was the best cut-off value that identify the risk of endometrial cancer.

Color Doppler score > 2 and Ill-defined endometrium- myometrium interface were the best ultrasound variables for predicting endometrial cancer with OR (51.80, 42) respectively and NPV (97.37, 97.30) respectively. Similar results were found by (N.M. Madkour., 2017 ; Stachowicz et al., 2021) [10,11].

Heterogeneous endometrial echogenicity was strongly associated with endometrial cancer (OR = 35.00, 95% CI: 4.279–400.7), with a sensitivity of 87.50% and specificity of 83.33%. However, its PPV of 50% suggests cautious interpretation of positive results. heterogeneous echogenicity and irregular endo-myometrial interface, observed in this study, were consistent with their predictive power reported in earlier research.[7,8]

Papadopoulos et al.[15] stated that; heterogeneous echogenicity of the endometrium and/or irregular endometrium-myometrium border further increases the risk.

The concept of using a scoring system to predict endometrial cancer is not novel, but many existing studies have faced significant limitations. These include overly complex models, impracticality, potential for biased outcomes, reliance on specialized equipment, or suboptimal diagnostic performance. For instance, Giannella et al. [9] developed the RHEA model, which incorporates factors such as recurrent vaginal bleeding, endometrial thickness exceeding 8 mm, hypertension, and age over 65 years. While this model demonstrated moderate diagnostic accuracy, it achieved an area under the curve (AUC) of 0.87, indicating reasonable predictive capability. Despite these promising results, challenges remain in refining such scoring systems for broader clinical applicability and improved diagnostic precision. In 2014 Dueholm et al, [16] developed the REC scoring system, incorporating BMI, Doppler score, endometrial thickness, interrupted endomyometrial junction, and irregular surface, demonstrated high accuracy with AUC to 0.95 (95% CI: 0.92–0.99), 91% sensitivity and 94% specificity at a REC score ≥4.

The proposed scoring model offers practical benefits, including improved risk stratification, reduction in unnecessary invasive procedures, and timely identification of high-risk patients. By facilitating early and accurate diagnosis, the model has the potential to significantly improve clinical outcomes and patient management strategies. The findings also highlight the need for incorporating standardized ultrasound terminology, such as those outlined by the International Endometrial Tumor Analysis (IETA) group, to enhance reproducibility and clinical adoption. While the study demonstrates promising results, its limitations include the relatively small sample size and single-center design, which may affect the generalizability of findings. Future research should focus on validating the scoring model across larger and more diverse populations and exploring additional parameters that could further refine diagnostic accuracy

## 6. Conclusion

The standardized terminology for describing endometrial ultrasound features, as proposed by the IETA group, offers significant clinical value by providing a consistent and reliable framework for endometrial assessment. Utilizing a malignancy risk model in cases of postmenopausal bleeding enables effective stratification of patients into low- and high-risk categories. By facilitating early detection and targeted intervention, this model has the potential to make a meaningful impact on patient care and outcomes in the field of gynecological oncology.

## Data Availability

All data produced in the present study are available upon reasonable request to the authors

## References

[1] Olatunde OA, Samaila MO, Imam MI, Uchime KE, Dauda SE. Histopathological patterns of endometrial carcinoma in a tertiary hospital in North-West Nigeria 2024. 10.3332/ecancer.2024.1651.

[2] Okunowo AA, Alakaloko MA, Ohazurike EO, Okunade KS, Anorlu RI. Trend and Characteristics of Endometrial Cancer in Lagos, Nigeria. Gulf J Oncolog 2019;1:52–9.

[3] Agarwal S, Melgandi W, Sonkar DR, Ansari FA, Arora S, Rathi AK, et al. Epidemiological characteristics of endometrial cancer patients treated at a tertiary health center in National Capital Territory of India. J Cancer Res Ther 2023;19:452. 10.4103/jcrt.jcrt_2029_21.

[4] Ring KL, Mills AM, Modesitt SC. Endometrial Hyperplasia. Obstet Gynecol 2022;140:1061– 75. 10.1097/AOG.0000000000004989.

[5] Cheney M, Shihab S, Vegunta S. Postmenopausal Bleeding: Assessment and Management. J Womens Health 2024;33:692–4. 10.1089/jwh.2023.0878.

[6] Braun MM. Diagnosis and Management of Endometrial Cancer. Endometrial Cancer 2016;93.

[7] Dueholm M, Hjorth IMD, Dahl K, Hansen ES, Ørtoft G. Ultrasound Scoring of Endometrial Pattern for Fast-track Identification or Exclusion of Endometrial Cancer in Women with Postmenopausal Bleeding. J Minim Invasive Gynecol 2019;26:516–25. 10.1016/j.jmig.2018.06.010.

[8] Schramm A, Ebner F, Bauer E, Janni W, Friebe-Hoffmann U, Pellegrino M, et al. Value of endometrial thickness assessed by transvaginal ultrasound for the prediction of endometrial cancer in patients with postmenopausal bleeding. Arch Gynecol Obstet 2017;296:319–26. 10.1007/s00404-017-4439-0.

[9] Giannella L, Mfuta K, Setti T, Cerami LB, Bergamini E, Boselli F. A Risk-Scoring Model for the Prediction of Endometrial Cancer among Symptomatic Postmenopausal Women with Endometrial Thickness > 4 mm. BioMed Res Int 2014;2014:1–7. 10.1155/2014/130569.

[10] Stachowicz N, Smolen A, Ciebiera M, Lozinski T, Poziemski P, Borowski D, et al. Risk Assessment of Endometrial Hyperplasia or Endometrial Cancer with Simplified Ultrasound-Based Scoring Systems. Diagnostics 2021;11:442. 10.3390/diagnostics11030442.

[11] Madkour N. An ultrasound risk-scoring model for prediction of endometrial cancer in post-menopausal women (using IETA terminology). Middle East Fertil Soc J 2017;22. 10.1016/j.mefs.2017.01.009.

[12] Seckin B, Cicek MN, Dikmen AU, Bostancı EI, Muftuoglu KH. Diagnostic value of sonography for detecting endometrial pathologies in postmenopausal women with and without bleeding. J Clin Ultrasound 2016;44:339–46. 10.1002/jcu.22329.

[13] Long B, Clarke MA, Morillo ADM, Wentzensen N, Bakkum-Gamez JN. Ultrasound detection of endometrial cancer in women with postmenopausal bleeding: Systematic review and meta-analysis. Gynecol Oncol 2020;157:624–33. 10.1016/j.ygyno.2020.01.032.

[14] Wong AS-W, Lao TT-H, Cheung CW, Yeung SW, Fan HL, Ng PS, et al. Reappraisal of endometrial thickness for the detection of endometrial cancer in postmenopausal bleeding: a retrospective cohort study. BJOG Int J Obstet Gynaecol 2016;123:439–46. 10.1111/1471-0528.13342.

[15] Papadopoulos V, Tsiveriotis K, Decavalas G. The Role of Ultrasound in Endometrial Cancer. Int J Clin Ther Diagn 2015:1–4. 10.19070/2332-2926-SI01001.

[16] Dueholm M, Møller C, Rydbjerg S, Hansen ES, Ørtoft G. An ultrasound algorithm for identification of endometrial cancer REC Score. Ultrasound Obstet Gynecol 2014;43:557–68. 10.1002/uog.13205.

